# Amiodarone for Atrial Fibrillation Cardioversion in Cardiac Surgery and Development of a Risk Prediction Model for Recurrence

**DOI:** 10.64898/2025.12.15.25342314

**Authors:** Linlin Chen, Qi Gao, Liangcheng Zhang

## Abstract

**Background:** The optimal timing and efficacy of pharmacological rhythm control in patients with preoperative atrial fibrillation (AF) undergoing cardiopulmonary bypass (CPB) surgery remain unclear. We aimed to evaluate the effectiveness of intravenous amiodarone administration during rewarming on early cardioversion and short-term outcomes, and to develop a predictive model for postoperative AF recurrence.

**Methods:** This retrospective cohort study included adult patients with preoperative atrial fibrillation who underwent cardiac surgery with CPB. Patients receiving a 150 mg intravenous bolus of amiodarone via the oxygenator at systemic rewarming initiation (nasopharyngeal temperature ≥32°C) during aortic cross-clamping (ACC) were defined as the amiodarone group (A group, n=423), and were compared with the non-intervention group (NI group, n=191). The primary outcome was sustained sinus rhythm within 12 hours post-cardioversion. Secondary outcomes included myocardial injury (measured by cardiac troponin I, cTnI), inflammatory response (neutrophil count, NEUT), renal function (blood urea nitrogen, BUN), use of inotropic support (milrinone, MIL) and renal replacement therapy (hemodialysis, HD), length of hospital stay (LOHS), and other short-term clinical endpoints. Inverse probability of treatment weighting (IPTW) was used to adjust for baseline imbalances. A multivariable logistic regression model was developed and validated to predict early AF recurrence.

**Results:** Of 614 patients, 423 were in the A group and 191 in the NI group. After IPTW adjustment, the A group had a significantly higher rate of sustained sinus rhythm (52.5% vs. 7.8%, *P*<0.001). They also exhibited lower levels of cTnI, NEUT, and BUN, reduced use of MIL and HD, and shorter LOHS, along with other favorable outcomes. NT-proBNP was transiently higher in the A group. The final prediction model—incorporating age, left atrial anteroposterior diameter (LAAD), left ventricular end-diastolic diameter (LVED), right ventricular anteroposterior diameter (RVAD), serum calcium, and comorbidity grade—showed strong discrimination (AUC: 0.866 in the training cohort, 0.795 in the validation cohort).

**Conclusion:** Amiodarone administration during rewarming improves early rhythm stability and short-term clinical outcomes in patients with preoperative AF undergoing CPB surgery. A validated risk prediction model identifies patients at high risk for recurrence, supporting individualized perioperative management strategies.

## 1. Introduction

Atrial fibrillation (AF) is the most prevalent sustained cardiac arrhythmia in clinical practice, with preoperative AF present in 12.6% to 19.2% of patients undergoing cardiac surgery^1–4^. AF causes particularly significant compromise to hemodynamic stability in cardiac surgery patients, as its rapid and disorganized atrial electrical activity results in loss of effective atrial contraction coupled with irregular ventricular rhythm, collectively reducing myocardial contractility and diminishing cardiac output by 20%-30%^5, 6^. Atrioventricular dyssynchrony additionally induces functional mitral regurgitation, further compromising stroke volume, while rapid ventricular contraction exacerbates myocardial oxygen consumption, potentially triggering an imbalance between oxygen supply and demand during the early reperfusion phase of cardiac recovery. Furthermore, shortened diastole and systolic blood pressure fluctuations impair coronary perfusion, delay myocardial functional recovery, and increase both postoperative inotropic drug requirements and dependency probability; studies demonstrate that preoperative atrial fibrillation correlates with elevated perioperative mortality and stroke risk in coronary artery bypass grafting (CABG) patients^7, 8^.

Amiodarone, a broad-spectrum class III antiarrhythmic agent, demonstrates well-documented efficacy for atrial fibrillation management through multifaceted pharmacological actions extending beyond potassium channel blockade to include mild sodium channel inhibition, noncompetitive α/β-adrenergic receptor antagonism, and calcium channel modulation^9^. Intravenous amiodarone achieved sinus rhythm conversion within 4 hours in 68.8% of non-surgical atrial fibrillation patients^10^. Among patients with persistent atrial fibrillation undergoing mitral valve replacement (MVR), intraoperative intravenous amiodarone achieved sinus rhythm restoration in 72.7% of cases within 72 hours after aortic cross-clamping (ACC) release^11^. However, its clinical benefits require further evaluation.

This study aims to investigate the cardioversion efficacy and short-term clinical outcomes of intravenous amiodarone administration during rewarming in cardiopulmonary bypass (CPB) surgery for patients with preoperative atrial fibrillation, while establishing and validating a risk prediction model for AF recurrence within 12 hours post-cardioversion to provide evidence-based rationale for proactive pharmacologic management of perioperative AF in cardiac surgery and optimize patient-specific dosage optimization strategies.

## 2. Methods

### 2.1 Study design

This retrospective cohort study received approval from the Institutional Review Board of Fuwai Shenzhen Hospital, Chinese Academy of Medical Sciences (IRB No. 2022107), with waiver of informed consent per institutional policy for observational research. The trial was registered at ClinicalTrials.gov (NCT06553391) on July 12, 2024. All procedures adhered to the STROBE guidelines (Strengthening the Reporting of Observational Studies in Epidemiology; version 2021).

### 2.2 Patient Selection

Electronic health records of 614 consecutive adults (≥18 years) undergoing on-pump cardiac surgery with documented preoperative persistent atrial fibrillation (defined per 2020 European Society of Cardiology [ESC] criteria) were retrospectively analyzed between June 2020 and December 2024.

Inclusion Criteria:

Documented persistent atrial fibrillation (per 2020 ESC criteria) confirmed by ≥2 preoperative electrocardiograms (12-lead or 24-hour Holter monitoring).

Scheduled for elective CABG and/or valve surgery. Exclusion Criteria:

Chronic amiodarone use (≥30 days preoperative administration).

Postoperative intravenous amiodarone initiated within 48 hours postoperatively. Paroxysmal atrial fibrillation (self-terminating episodes <7 days).

Emergency surgical procedures.

Prior catheter-based atrial fibrillation ablation.

Use of other antiarrhythmic drugs (e.g., β-blockers, propafenone, flecainide, sotalol) within 48 hours prior to surgery.

### 2.3 Administration Protocol

Patients were assigned to two groups based on intraoperative management strategies. The Amiodarone Group (A Group, n=423), who received a 150 mg intravenous amiodarone bolus via the CPB oxygenator at systemic rewarming initiation (nasopharyngeal temperature ≥32°C) during ACC. The Non-Intervention Group (NI Group, n=191) received standard intraoperative care without proactive antiarrhythmic or hemodynamic support. The selection and dosing of anesthetic induction and maintenance agents lack standardized protocols, relying entirely on anesthesiologists’ clinical discretion. Rescue medications (e.g., atropine, phenylephrine) were administered at anesthesia team discretion for life-threatening instability per institutional protocols.

### 2.4 Data collection

Patient data were extracted from the DoCare Anesthesia Clinical Information System (Suzhou Mediastone Medical Technology Co., Ltd., Version 5.0, 2024) and categorized into four primary domains: (1) basic demographic and clinical information; (2) preoperative cardiac function and laboratory indicators; (3) perioperative surgical and medication-related indicators; and (4) postoperative recovery and outcome indicators.

The first domain included age, height, weight, primary diagnosis, medical history—with a focus on atrial fibrillation—and preoperative medication use, specifically beta-blockers and digoxin. The second domain encompassed preoperative cardiac structural and functional parameters: left atrial anteroposterior diameter (LAAD), left ventricular end-diastolic diameter (LVED), right atrial left-right diameter (RAM), right ventricular anteroposterior diameter (RVAD), interventricular septum thickness (IVS), left ventricular ejection fraction (EF), and pulmonary arterial pressure (PAP). Laboratory assessments included glycated hemoglobin A1c (HbA1c), N-terminal pro-B-type natriuretic peptide (NT-proBNP), cardiac troponin I (cTnI), cardiac troponin T (cTnT), creatine kinase-MB (CK-MB), serum creatinine (Cr), estimated glomerular filtration rate (eGFR), blood urea nitrogen (BUN), white blood cell count (WBC), neutrophil percentage (NEUT), and C-reactive protein (CRP).

Perioperative indicators (third domain) included surgical and anesthesia variables such as type of operation, anesthesia duration (An time), cardiopulmonary bypass time (CPB time), aortic cross-clamp (ACC) release time, status of spontaneous cardiac rewarming (Rerouting), heart rhythm after rewarming (HR), timing of amiodarone administration, time to sinus rhythm restoration (Sinus rhythm), time from rewarming to sinus rhythm recovery (TRSR), duration of sustained sinus rhythm maintenance (TSRM), and date of atrial fibrillation recurrence (DAFR). Additionally, perioperative use of vasoactive and inotropic agents—including dopamine (DA), dobutamine (DOB), epinephrine (AE), norepinephrine (NE), milrinone (MIL), and vasopressin—as well as electrolyte and metabolic parameters (serum Na, K, Ca, Mg, glucose [Glu], and lactate [Lac]) measured within 1 hour after CPB termination were recorded.

The primary endpoint was cardioversion success rate, defined as sustained sinus rhythm without recurrence of atrial fibrillation from aortic cross-clamp (ACC) release up to and including the 12-hour postoperative time point. Sustained sinus rhythm was confirmed by the presence of ≥3 consecutive sinus P-waves on continuous 5-lead electrocardiographic (ECG) monitoring, with rhythm assessments performed at least every 15 minutes during the intraoperative period and continuously in the intensive care unit (ICU). Any documented episode of atrial fibrillation lasting ≥30 seconds during this interval was considered a recurrence and classified as cardioversion failure.

Secondary outcomes included both clinical efficacy and safety measures: spontaneous conversion to sinus rhythm, need for temporary pacing, duration of sustained regular rhythm, use of vasoactive medications, unplanned reoperation (Secondary), requirement for postoperative hemodialysis (HD), ICU length of stay (LOICU), duration of mechanical ventilation, and hospital length of stay (LOHS). Postoperative laboratory evaluations included serial measurements of NT-proBNP, cTnI, Cr, BUN, WBC, and NEUT on postoperative days 1–3. These composite endpoints enabled a comprehensive assessment of the efficacy and safety of the compared treatment regimens in patients undergoing cardiac surgery with preoperative persistent atrial fibrillation.

All data were independently extracted by two researchers, with discrepancies resolved through consensus adjudication by a third senior cardiac surgeon to ensure accuracy and reliability. Additionally, all data were fully anonymized prior to analysis; researchers had no access to identifiable patient information throughout the process, thereby protecting the privacy of study participants while maintaining the integrity of the dataset.

### 2.5 Statistical analysis

To minimize selection bias and balance baseline confounding factors between treatment groups, inverse probability of treatment weighting (IPTW) based on propensity scores (PS) was employed to create a pseudo-population in which observed covariates were balanced across groups. The propensity score was estimated using a logistic regression model predicting treatment assignment from baseline characteristics. To enhance the stability and robustness of the IPTW estimator, extreme weights were truncated at the 99th percentile (i.e., weight trimming), thereby reducing the influence of high-leverage observations that could distort effect estimates.

Continuous variables were summarized as mean ± standard deviation (SD) if normally distributed, and compared between groups using the independent samples *t*-test; non-normally distributed continuous variables were presented as median (interquartile range: 25th–75th percentiles) and analyzed using the Mann–Whitney *U* test. Categorical variables were expressed as frequency and percentage [n (%)] and compared using the chi-square test or Fisher’s exact test, as appropriate.

All eligible patients were randomly divided into a training set and a validation set in an 8:2 ratio using simple random sampling. Stratification was applied during the split to preserve the proportion of atrial fibrillation recurrence outcomes in both sets, minimizing potential selection bias.

Within the training cohort, univariate logistic regression analyses were performed for all candidate covariates to identify potential predictors of postoperative atrial fibrillation recurrence. Variables with a univariate *p*-value < 0.2, along with clinically relevant predefined variables, were retained for inclusion in the multivariable logistic regression model. This final model was used to estimate the probability of successful cardioversion—defined as sustained sinus rhythm within 12 hours postoperatively—adjusting for potential confounders. Odds ratios (ORs) with corresponding 95% confidence intervals (CIs) were reported.

Model discrimination was assessed by calculating the area under the receiver operating characteristic curve (AUC) separately in both the training and validation cohorts, and ROC curves were plotted accordingly. To facilitate clinical interpretation, a graphical nomogram or bar chart was developed to visualize the contribution of each predictor variable in the model and the predicted probability of atrial fibrillation recurrence based on a total risk score.

## 3. Result

### 3.1 Patient characteristics

This study included 614 patients assigned to Group NI (n = 191) and Group A (n = 423). To correct potential baseline imbalances, we applied the Inverse Probability of Treatment Weighting (IPTW) method based on propensity scores (PS) and truncated extreme weights at the 99th percentile. The following presents a comparison of demographic characteristics before and after the adjustment (Table 1).

**Table 1.**
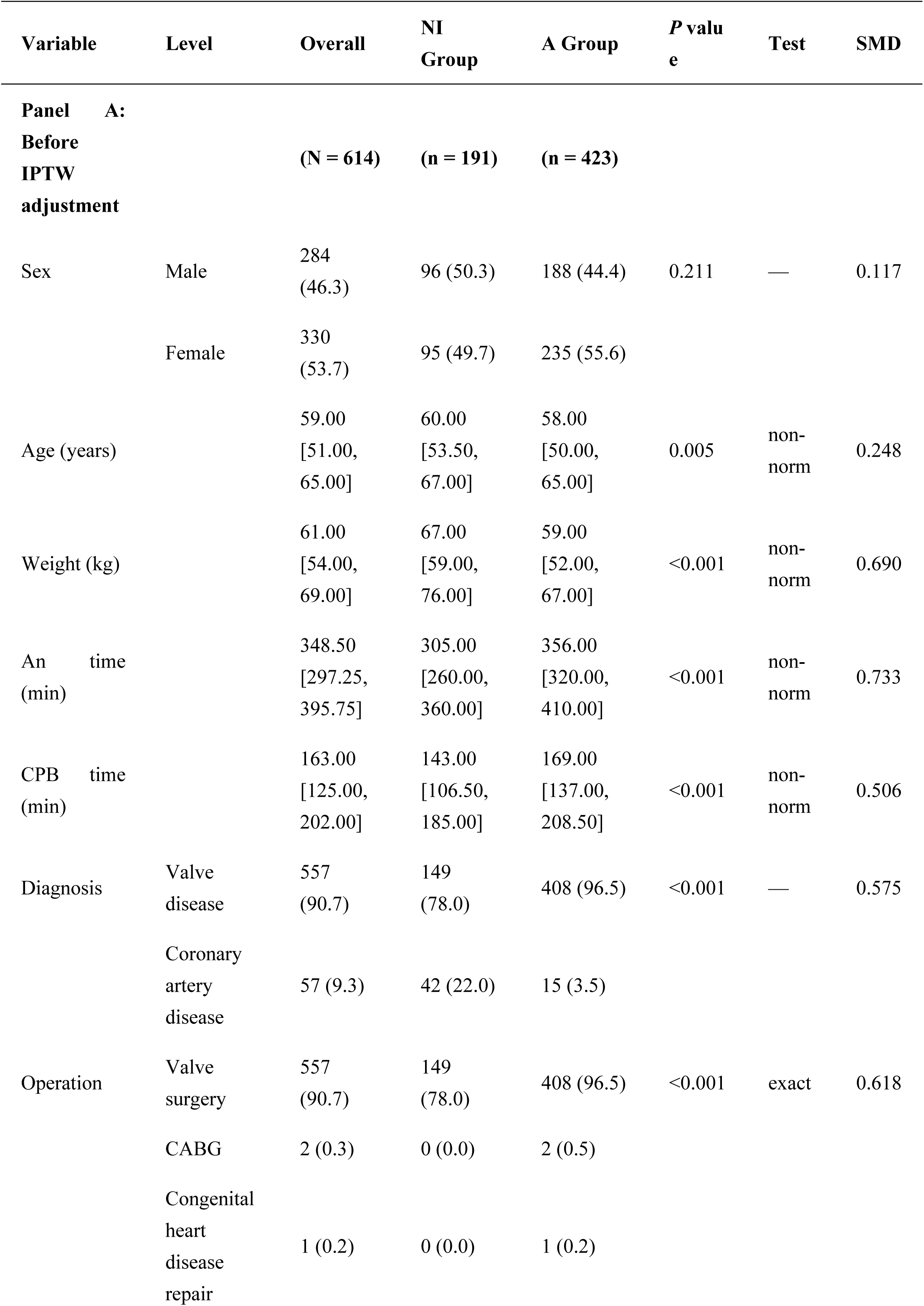
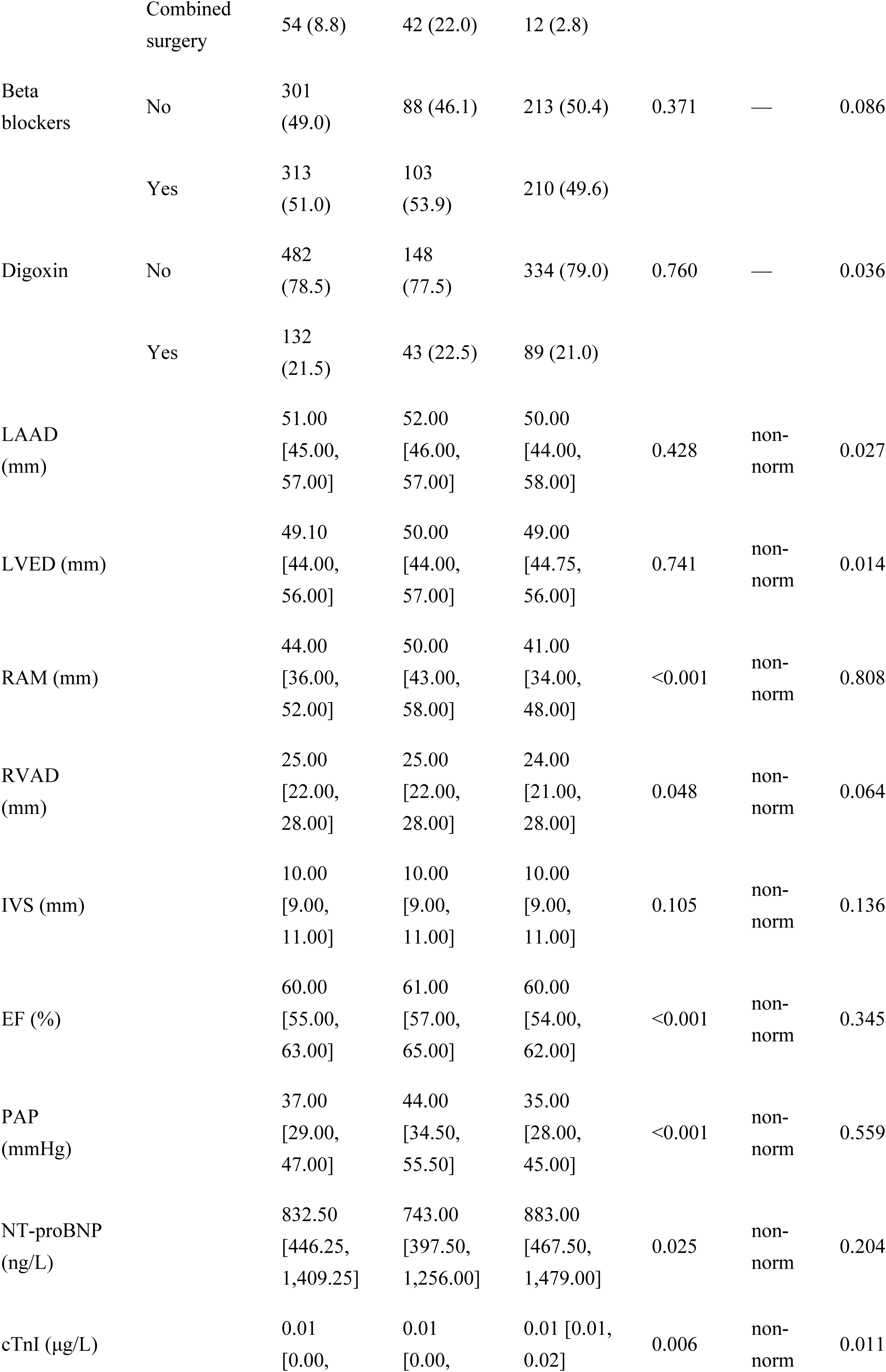
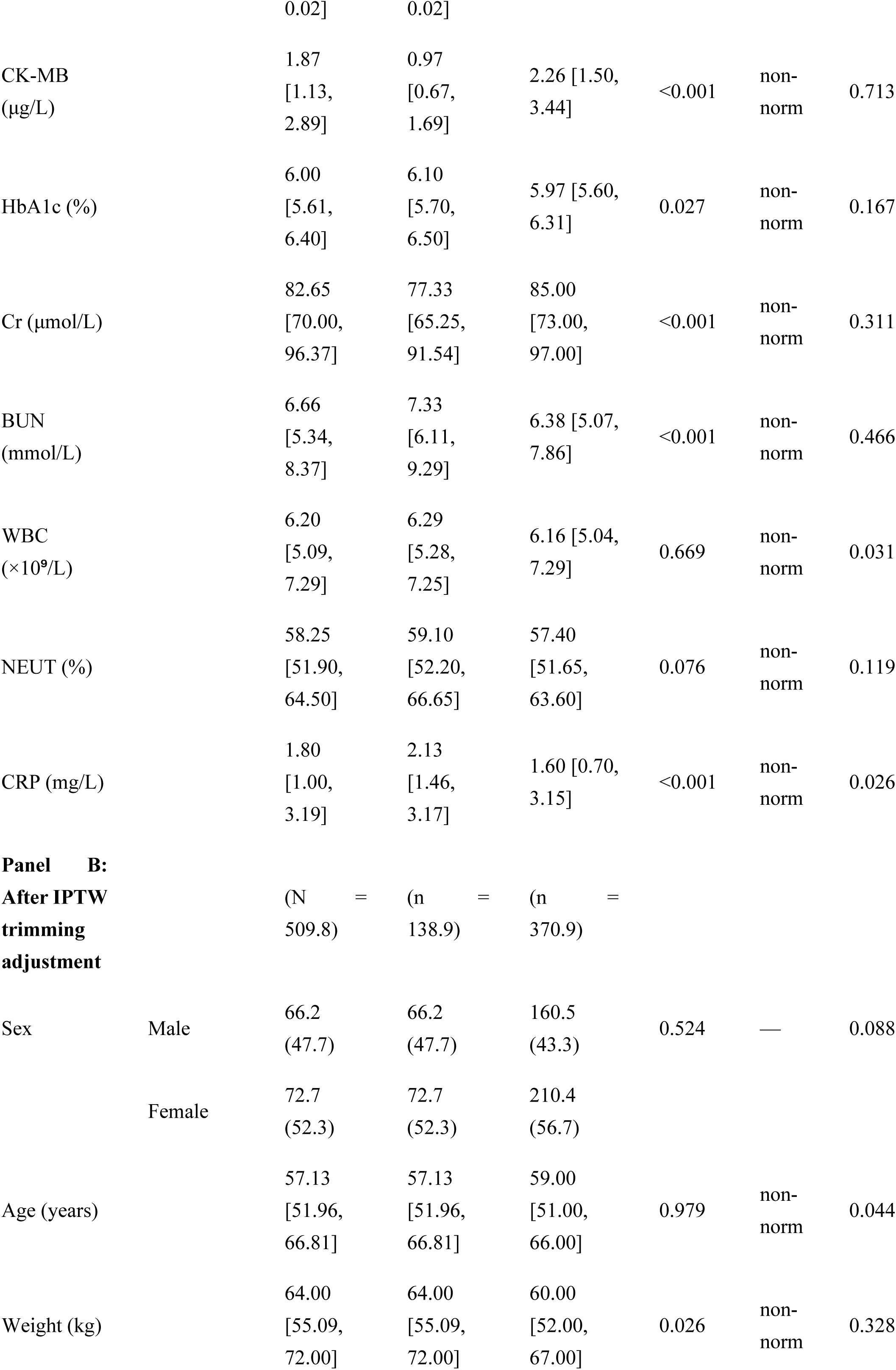
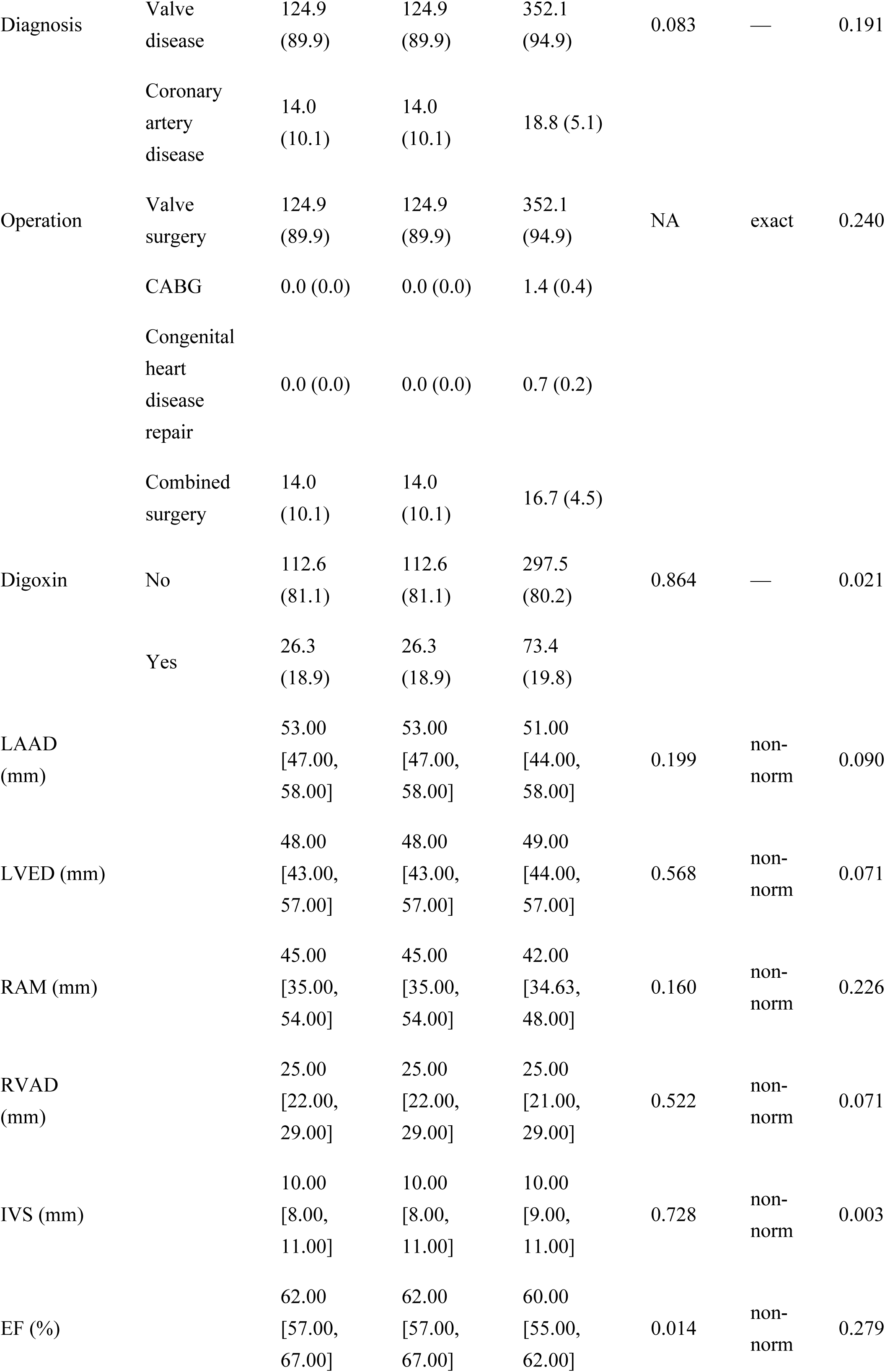
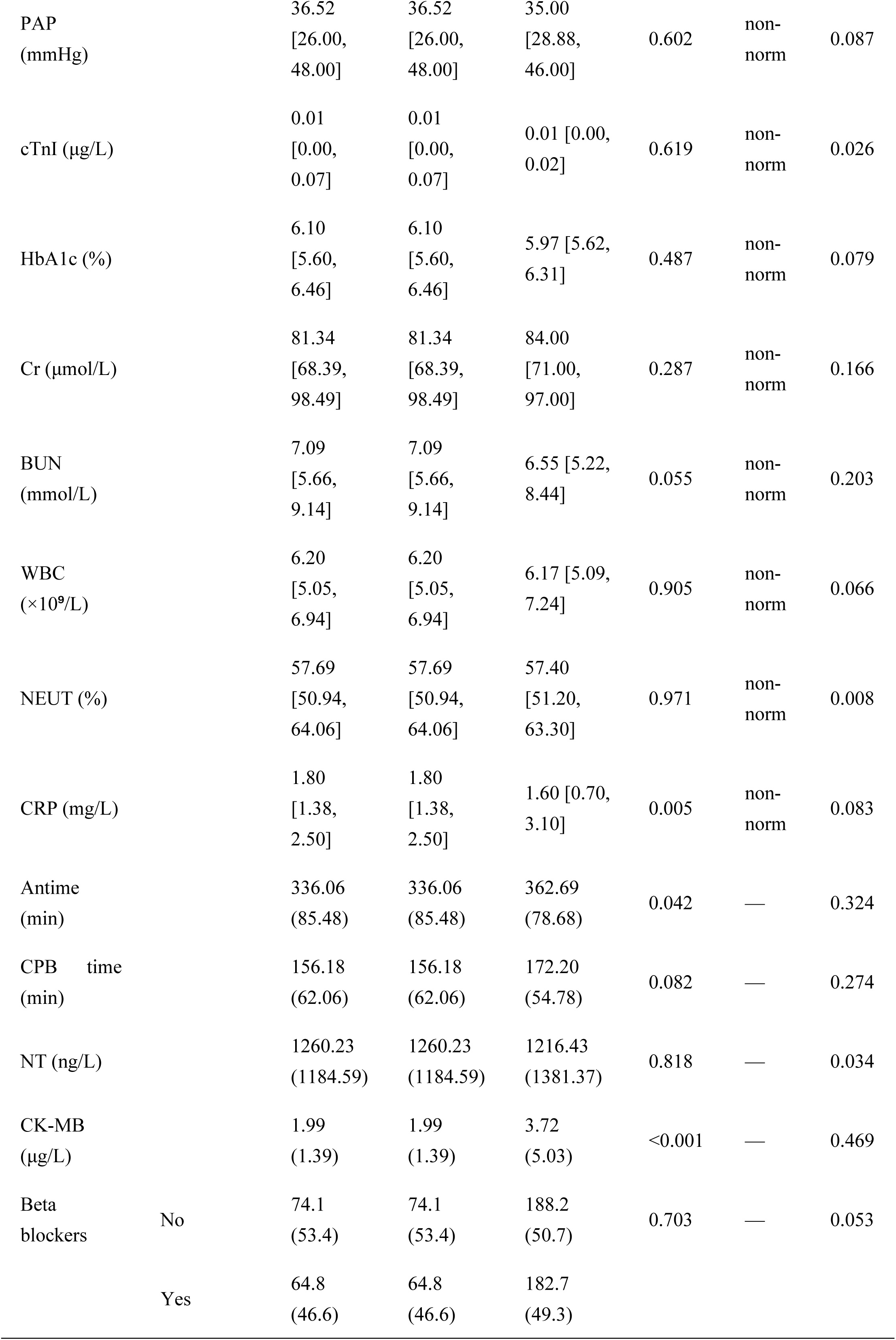
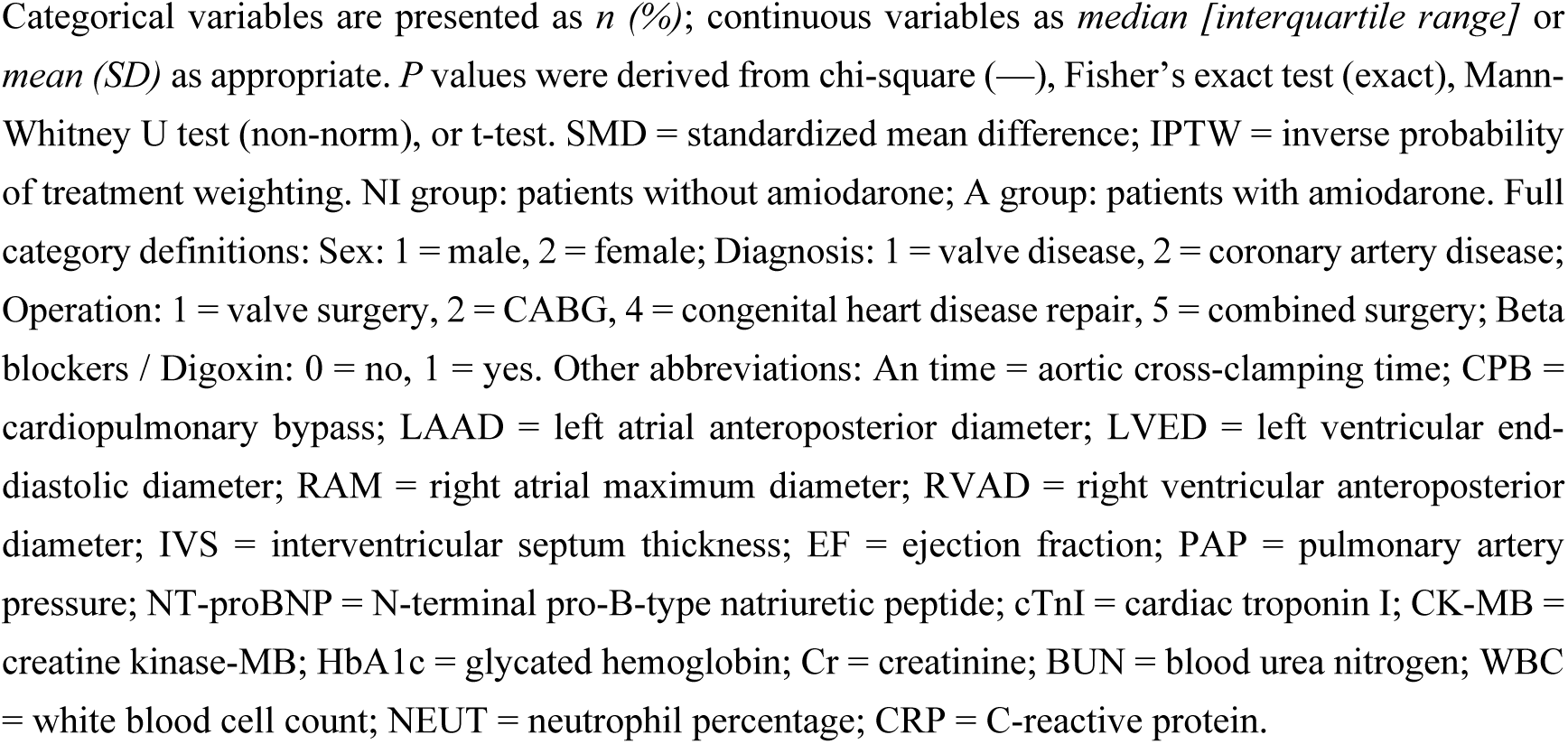
Patient Characteristics Before and After Inverse Probability of Treatment Weighting with Trimming.

Before adjustment, there were significant statistical differences between the two groups in multiple indicators. In terms of basic characteristics, the median age in Group NI was 60.00 [53.50, 67.00] years, while in Group A it was 58.00 [50.00, 65.00] years (p = 0.005). The median weight in Group NI was 67.00 [59.00, 76.00] kg, compared to 59.00 [52.00, 67.00] kg in Group A (p < 0.001). Regarding treatment - related indicators, the median anesthesia time in Group NI was 305.00 [260.00, 360.00] minutes, while in Group A it was 356.00 [320.00, 410.00] minutes (p < 0.001). The median cardiopulmonary bypass time in Group NI was 143.00 [106.50, 185.00] minutes, and in Group A it was 169.00 [137.00, 208.50] minutes (p < 0.001). For the distribution of diagnosis categories and surgical methods, the proportion of a certain diagnosis category was 78.0% in Group NI and 96.5% in Group A (p < 0.001). In terms of drug use, the proportion of beta - blocker use was 46.1% in Group NI and 50.4% in Group A (p = 0.371), and for digoxin use, it was 77.5% in Group NI and 79.0% in Group A (p = 0.760). There were significant differences in cardiac indicators. The median left - right diameter of the right atrium in Group NI was 50.00 [43.00, 58.00] mm, while in Group A it was 41.00 [34.00, 48.00] mm (p < 0.001). The median pulmonary arterial pressure in Group NI was 44.00 [34.50, 55.50] mmHg, and in Group A it was 35.00 [28.00, 45.00] mmHg (p < 0.001). Among laboratory indicators, there were significant differences between the two groups in indicators such as creatine kinase - MB, serum creatinine, and blood urea nitrogen (all p - values < 0.001).

After the adjustment using the IPTW method with weight truncation, there were no statistical differences between the two groups in all of the above - mentioned indicators. Overall, the IPTW method combined with weight truncation effectively balanced the differences between the two groups in multiple indicators, making the two groups more comparable.

### 3.2 Screening of Prognosis-Related Factors

Through univariate Logistic regression analysis, a total of 17 factors related to the prognosis of patients after cardiac surgery were screened out, including Sex, Age, medical history, HR, beta, LAAD, LVED, RAM, RVAD, IVS, PAP, Na, Ca, Glu, NT, Cr, and BUN. Combined with clinical practice, 9 clinical prognosis-related variables were further screened, namely Age, medical history, beta, Digoxin, LAAD, EF, PAP, and CRP.

### 3.3 Primary and Secondary Outcome Measures

The primary outcome of successful cardioversion was significantly higher in the A group compared to the NI group, with rates of 52.5% versus 7.8%, respectively (*P* < 0.001) (Table 2), indicating a substantial benefit of the intervention in restoring sinus rhythm.

**Table 2.**
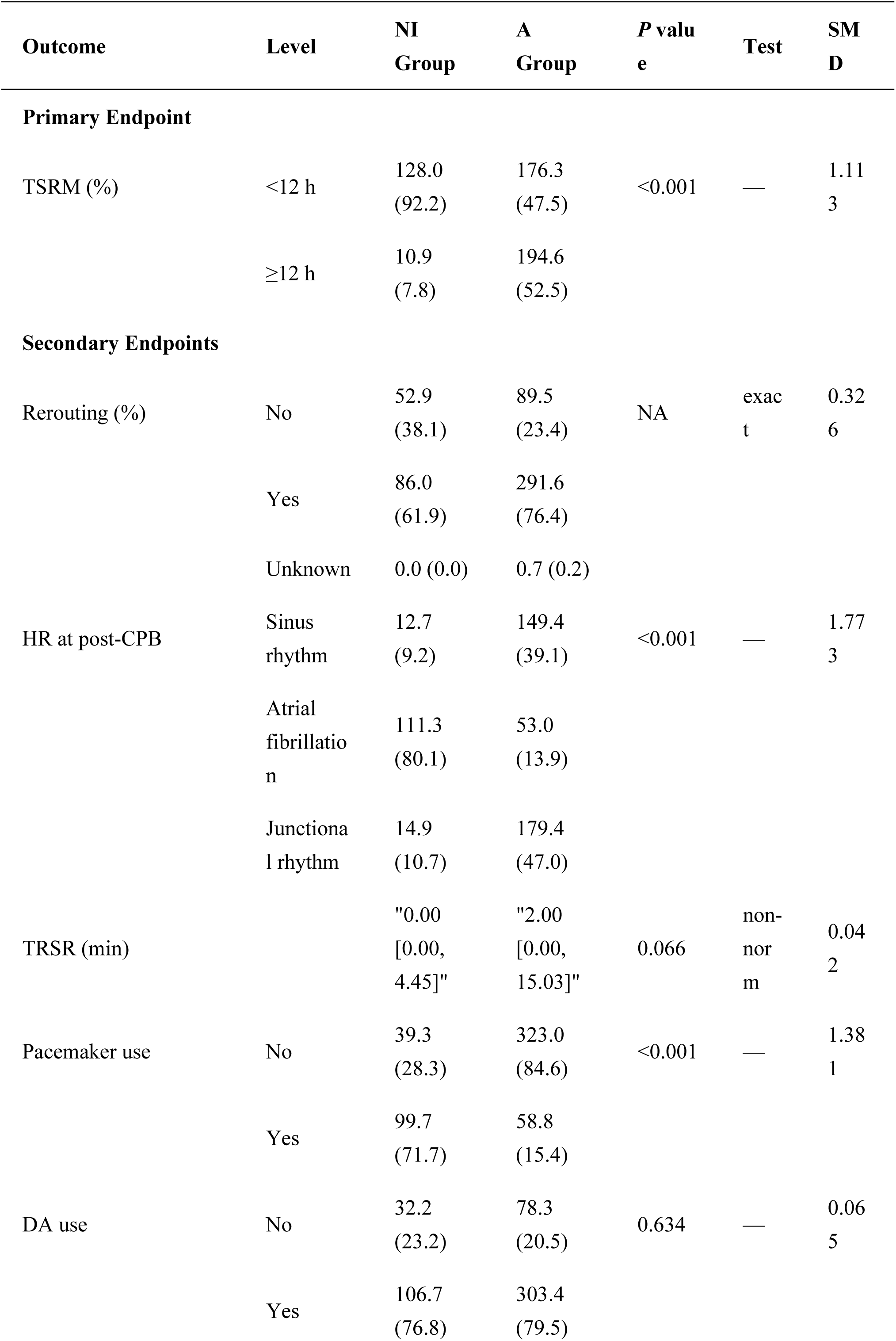
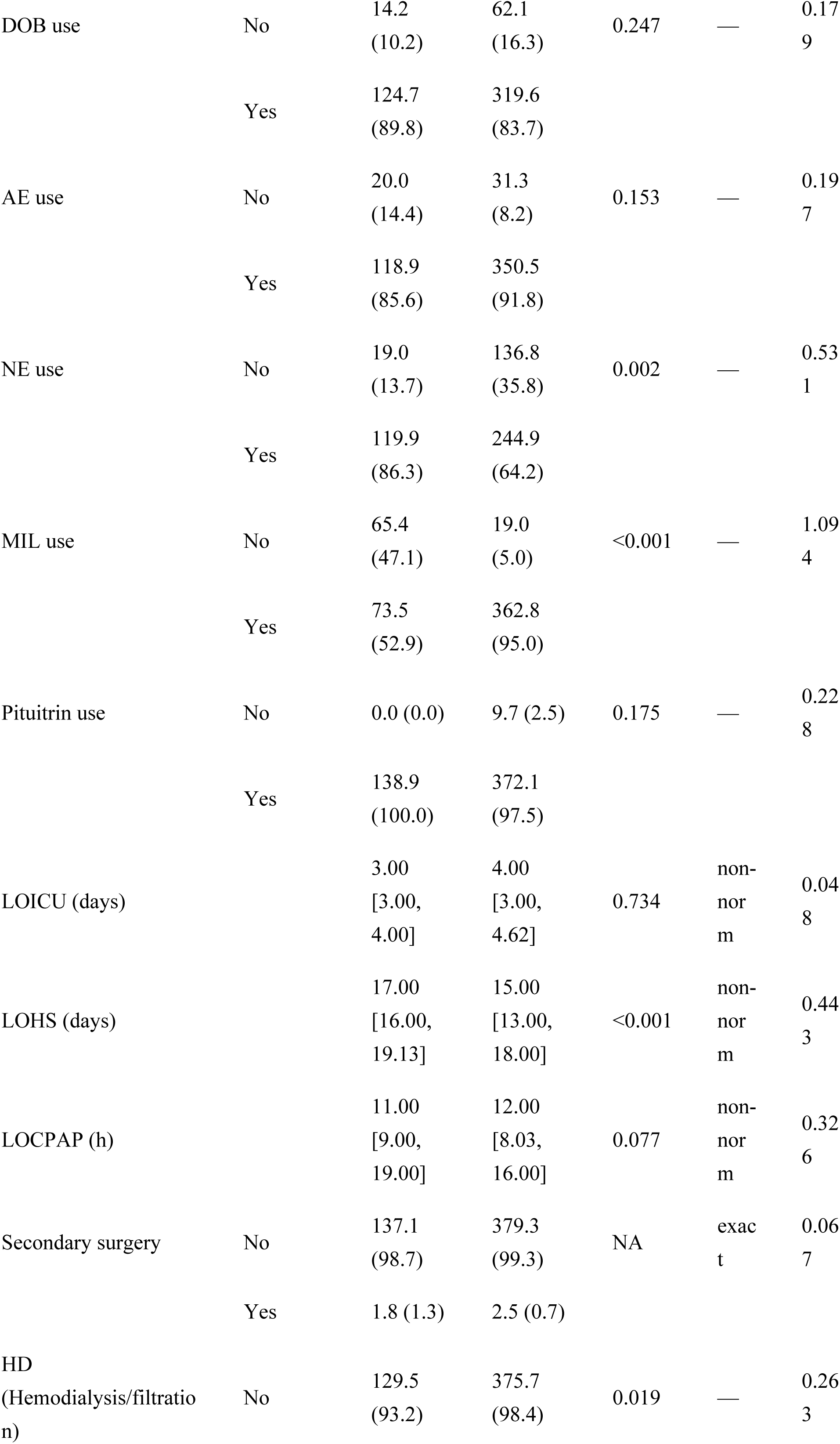
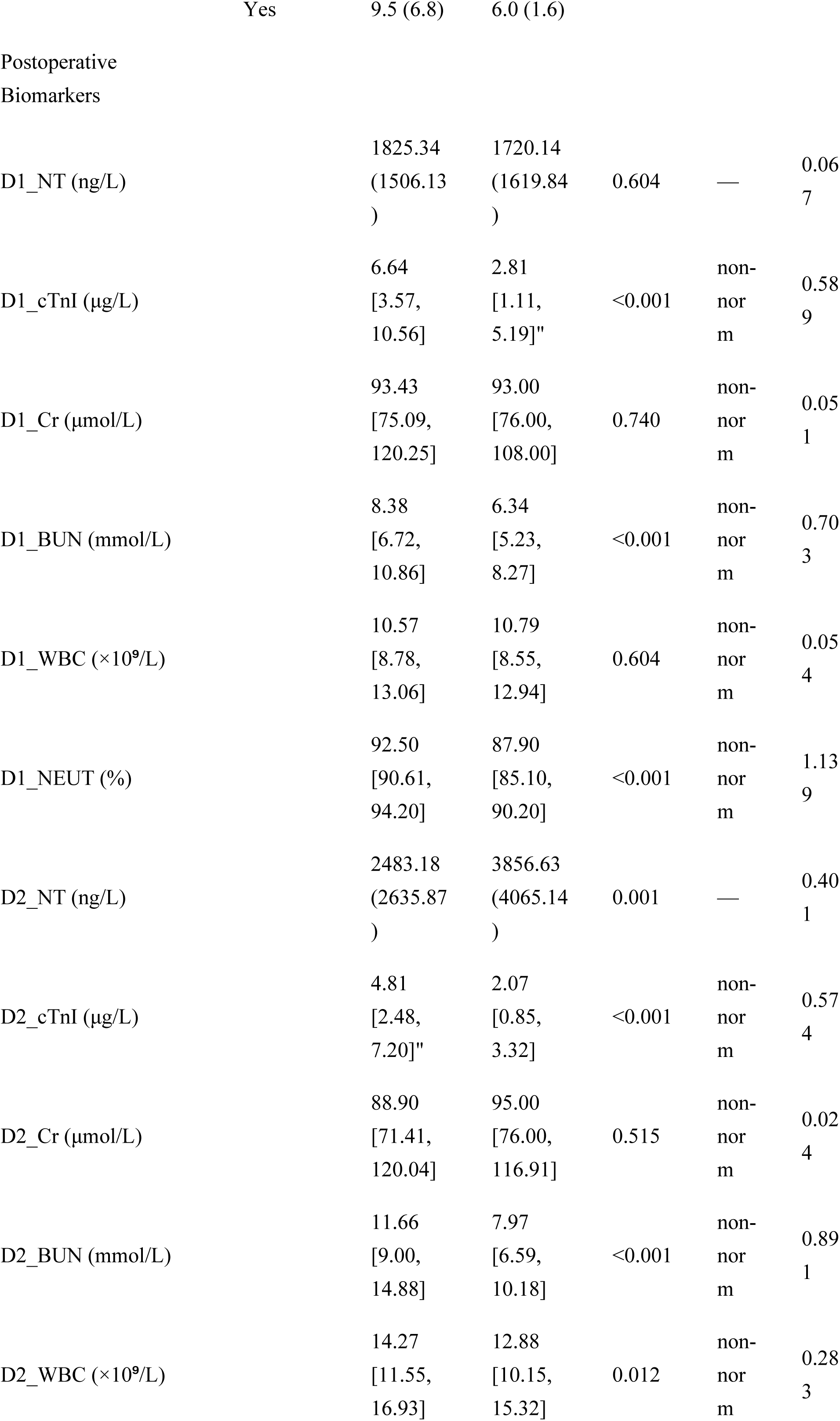
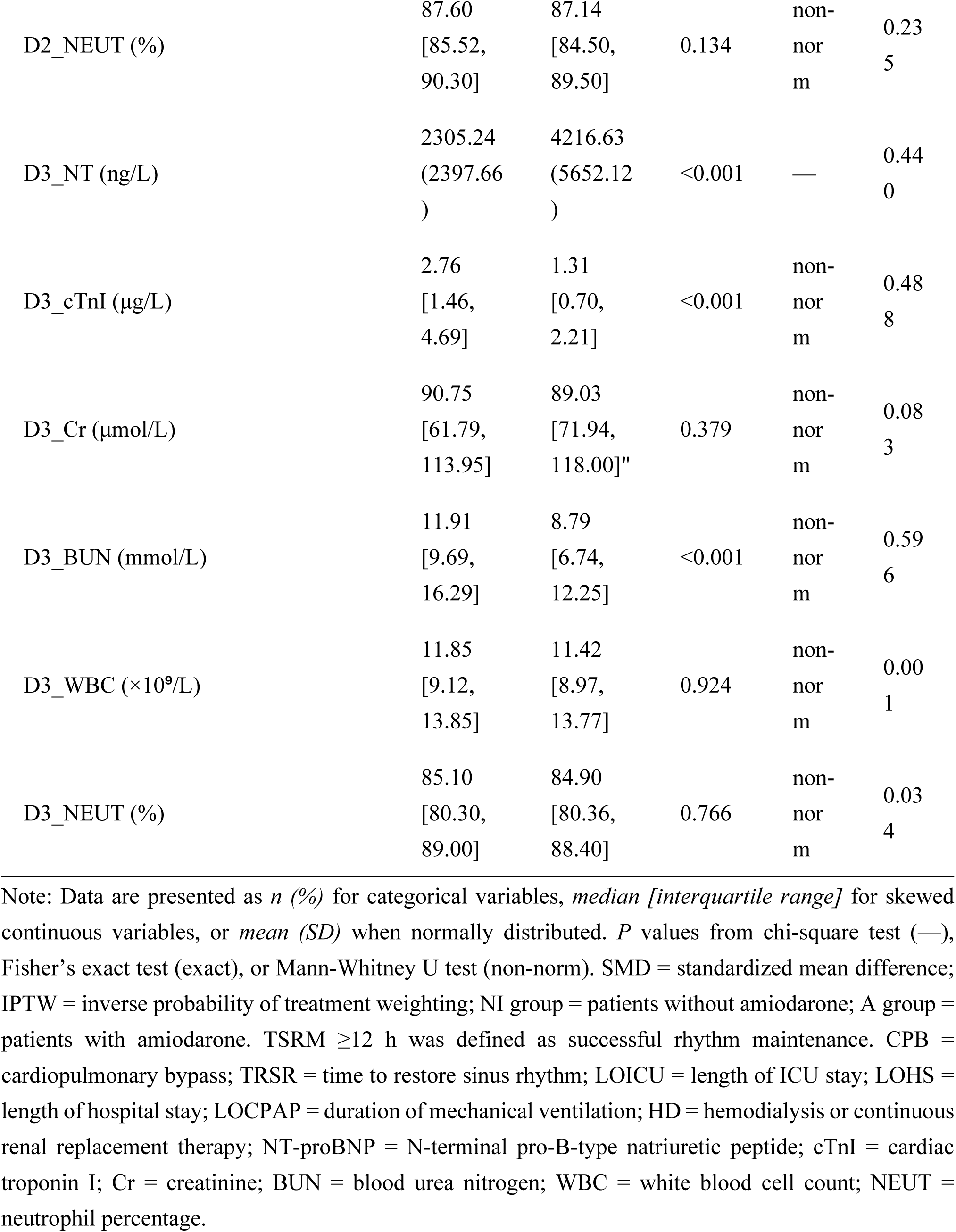
Primary and Secondary Outcomes in the Inverse Probability of Treatment Weighting (IPTW)-Trimmed Population.

Among secondary outcomes (Table 2), the NI group exhibited greater myocardial injury, as reflected by significantly elevated postoperative cTnI levels at all measured time points: D1_cTnI (6.64 vs. 2.81 ng/mL, *P* < 0.001), D2_cTnI (4.81 vs. 2.07 ng/mL, *P* < 0.001), and D3_cTnI (2.76 vs. 1.31 ng/mL, *P* < 0.001). In contrast, NT-proBNP levels were significantly higher in the A group on postoperative day 2 (D2_NT: 2483.18 vs. 3856.63 pg/mL, *P* = 0.001) and day 3 (D3_NT: 2305.24 vs. 4216.63 pg/mL, *P* < 0.001), suggesting that despite lower initial myocardial injury, patients in the A group experienced greater ventricular wall stress or neurohormonal activation during later recovery (Figure 1).

**Figure 1.**
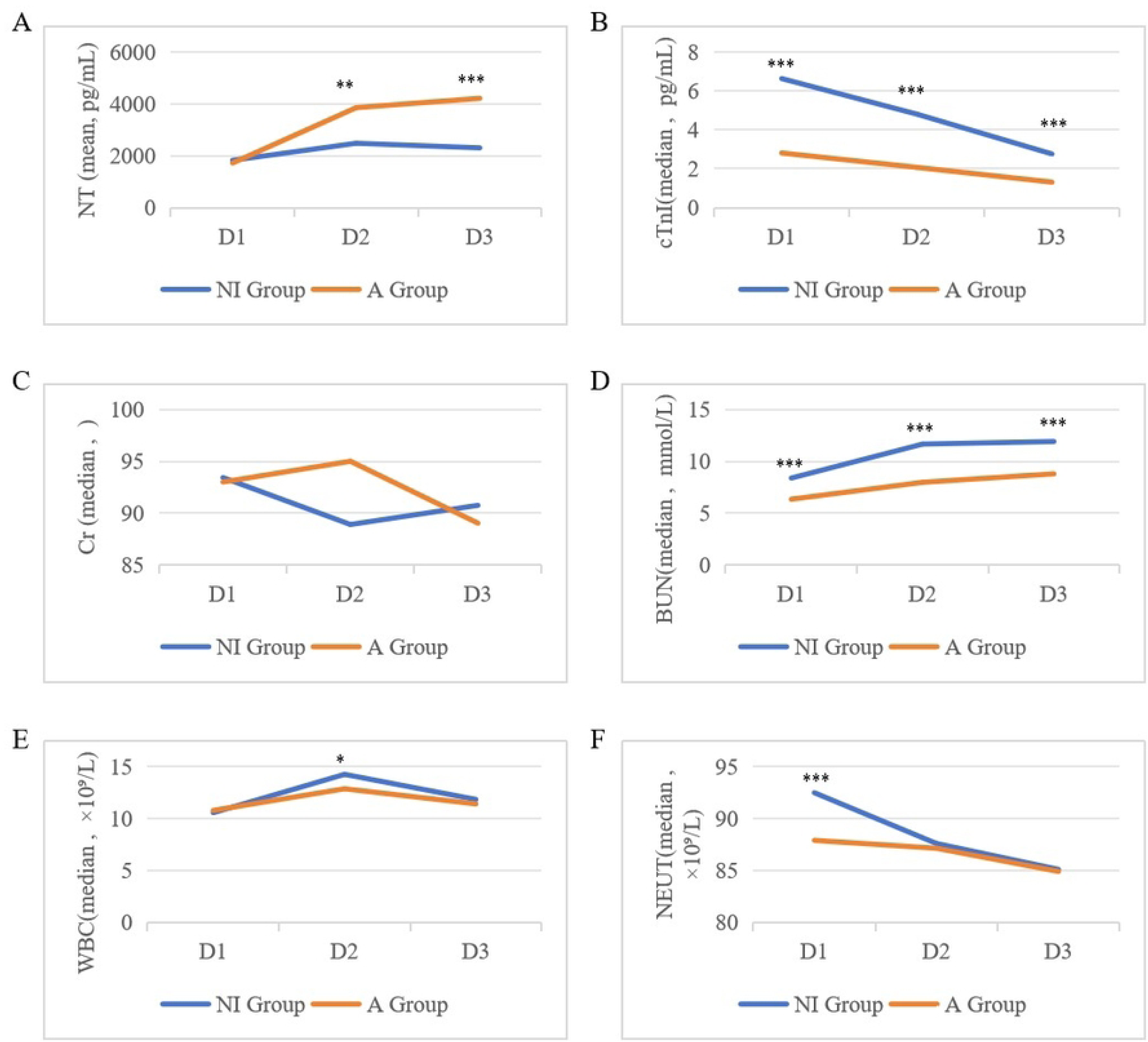
Line Graph Showing Variations in Laboratory Indicators of the NI Group vs. A Group From Postoperative Day 1 to Day 3. A: NT; B: cTnI; C: Cr; D: BUN; E: WBC; F: NEUT. NT = N-terminal pro-B-type natriuretic peptide; cTnI = Cardiac troponin I; Cr = creatinine level; BUN = blood urea nitrogen; WBC = White blood cell count; NEUT = Neutrophil count. \**P*<0.05; **P<0.01; \*\*\**P* <0.001

Pharmacological support differed markedly between groups. MIL use was far more common in the NI group (47.1% vs. 5.0%, *P* < 0.001), whereas NE was used more frequently in the A group (13.7% vs. 35.8%, *P* < 0.001). There were no significant differences in the use of DA (*P* = 0.634), DOB (*P* = 0.247), AE (*P* = 0.153), or pituitrin (*P* = 0.175). HD was more commonly performed in the NI group (6.8% vs. 1.6%, *P* = 0.019), indicating greater need for renal replacement therapy, although serum Cr levels (D1_Cr, D2_Cr, D3_Cr) did not differ significantly between groups (*P* > 0.05 for all). The systemic inflammatory response was stronger in the NI group in the early postoperative phase. NEUT on postoperative day 1 (D1_NEUT) was significantly higher in the NI group (92.50% vs. 87.90%, *P* < 0.001), with a large effect size (SMD = 1.139). WBC on day 2 (D2_WBC) was also higher in the NI group (14.27 vs. 12.88 ×10⁹/L, *P* = 0.012), but this difference was not sustained on day 3 (*P* = 0.924). No significant differences were observed in D2_NEUT (*P* = 0.134) or D3_NEUT (*P* = 0.766).

Metabolic markers showed consistent patterns: BUN was significantly higher in the NI group at all postoperative time points — D1_BUN (8.38 vs. 6.34 mmol/L, *P* < 0.001), D2_BUN (11.66 vs. 7.97 mmol/L, *P* < 0.001), and D3_BUN (11.91 vs. 8.79 mmol/L, *P* < 0.001) — suggesting greater catabolic stress or fluid imbalance. A more direct change can be seen in Figure 1.

In terms of clinical recovery, the NI group had a significantly longer LOHS (17.00 vs. 15.00 days, *P* < 0.001). No significant differences were observed in LOICU, LOCPAP, or incidence of Secondary.

### 3.4 Results of Multivariate Logistic Regression Analysis

Multivariate Logistic regression analysis was conducted to clarify the predictive effect of various indicators on the probability of patients recovering and maintaining sinus rhythm for 12 hours after cardiac surgery. The results showed that Age, LAAD, LVED, RVAD, Ca, and medical history (Grade 3 vs. Grade 1) were significant predictive factors for this outcome (all P<0.05), while 14 indicators including EF, RAM, and PAP had no statistical significance on the outcome (all *P*>0.05). The detailed results are as follows:

### Predictive Factors with Significant Impact on Outcome

Age had a negative predictive effect on the recovery and maintenance of sinus rhythm for 12 hours after surgery (OR=0.478, 95%CI: 0.288–0.792, *P*<0.05), meaning that for each 1-year increase in age, the probability of patients recovering and maintaining sinus rhythm after surgery decreased significantly, with the odds ratio decreasing by approximately 52%. LAAD was negatively correlated with the outcome (OR=0.520, 95%CI: 0.327–0.827, *P*<0.05); for each 1-unit increase in LAAD, the probability of patients recovering sinus rhythm decreased significantly, with the odds ratio decreasing by approximately 48%. LVED was a negative predictive factor for the outcome (OR=0.572, 95%CI: 0.385–0.851, *P*<0.05); for each 1-unit increase in LVED, the probability of patients recovering and maintaining sinus rhythm for 12 hours after surgery decreased significantly, with the odds ratio decreasing by approximately 43%. There was a negative correlation between RVAD and the outcome (OR=0.667, 95%CI: 0.488–0.911, *P*<0.05); for each 1-unit increase in RVAD, the probability of patients recovering and maintaining sinus rhythm decreased significantly. Ca had a negative predictive effect on the outcome (OR=0.724, 95%CI: 0.548–0.955, *P*<0.05); for each 1-unit increase in Ca concentration, the probability of patients recovering sinus rhythm after surgery decreased significantly, with the odds ratio decreasing by approximately 28%. Compared with patients with medical history=Grade 1, patients with medical history=Grade 3 had a significantly lower probability of recovering and maintaining sinus rhythm for 12 hours after surgery (OR=0.119, 95%CI: 0.048–0.294, *P*<0.05), with the odds ratio decreasing by approximately 88%.

### Variables with No Significant Impact on Outcome

EF had no significant predictive effect on the outcome (OR=0.963, 95%CI: 0.741–1.253, *P*>0.05), with an effect size of -0.037. RAM had no significant association with the outcome (OR=0.941, 95%CI: 0.615–1.439, *P*>0.05). Although PAP showed a negative trend with the outcome, it did not reach statistical significance (OR=0.680, 95%CI: 0.457–1.012, *P*>0.05). In addition, CRP, IVS, Na, Glu, NT, Cr, BUN, medical history (Grade 2 vs. Grade 1), beta, HR, and Digoxin had no significant impact on the probability of patients recovering and maintaining sinus rhythm for 12 hours after cardiac surgery (all *P*>0.05).

### 3.5 Evaluation of Predictive Model Performance

The area under the receiver operating characteristic curve (AUC) of the predictive model constructed based on multivariate Logistic regression analysis was 0.866 (95%CI: 0.829–0.904) in the training set and 0.795 (95%CI: 0.695–0.894) in the validation set (Figure 2). This indicates that the model has good predictive performance and discriminative ability, and can effectively identify high-risk and low-risk patients who recover and maintain sinus rhythm for 12 hours after cardiac surgery. To improve the clinical applicability of the model, it has been converted into a nomogram (Figure 3), which can quickly estimate the probability of patient outcomes through an intuitive scoring method.

**Figure 2.**
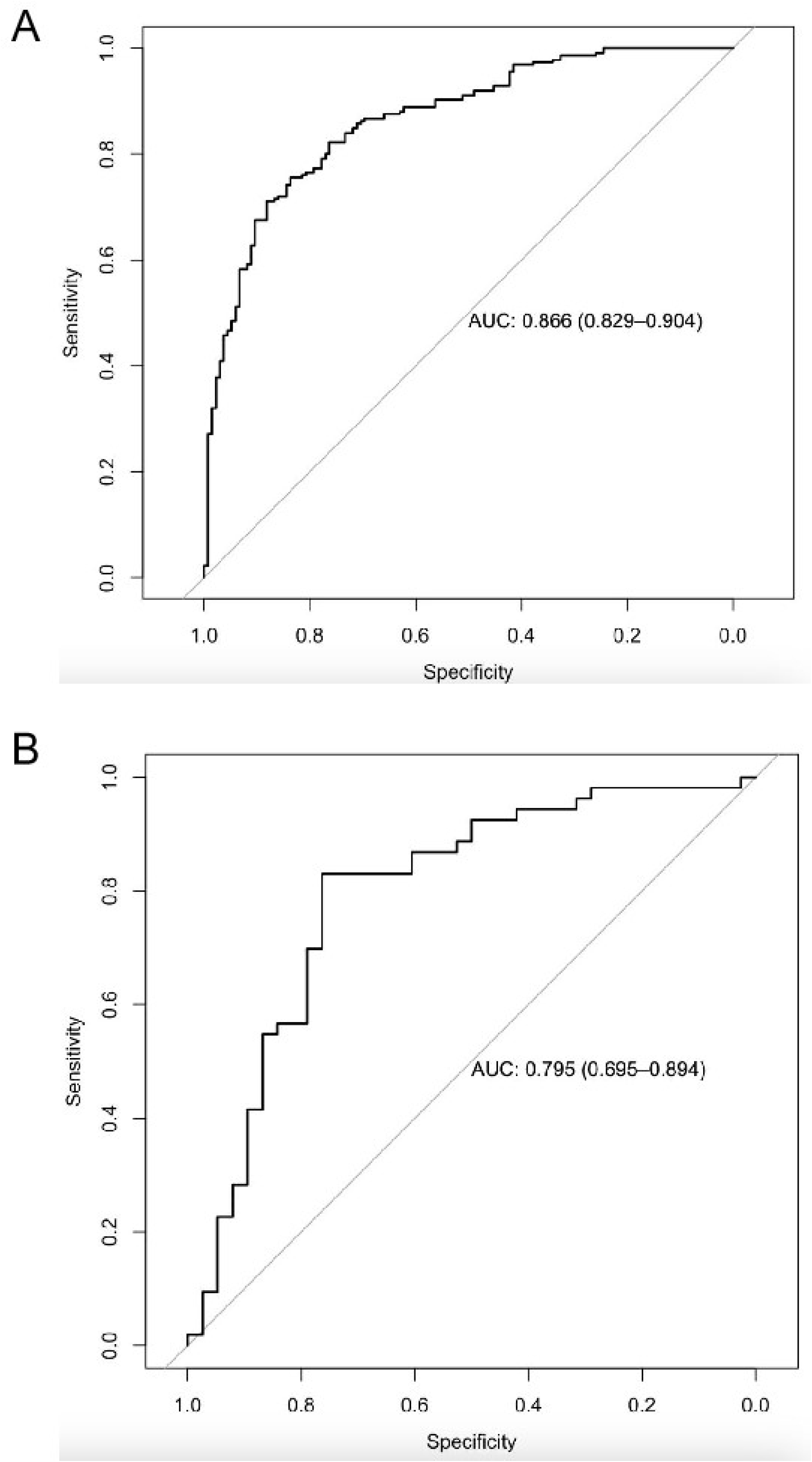
AUC curves of training and validation sets. A: The training set; B: The validation set.

**Figure 3.**
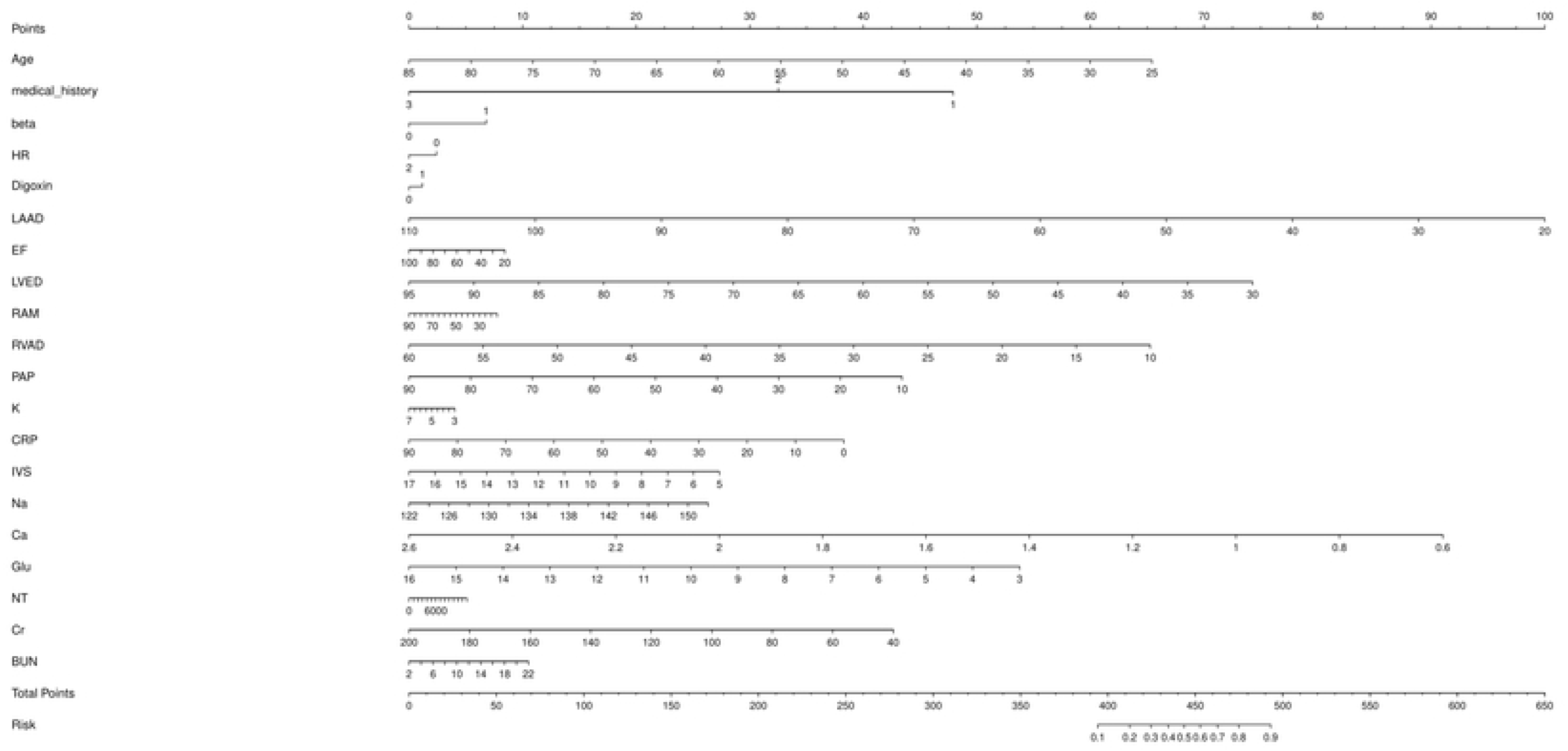
Nomogram for predicting the probability of restoring and maintaining sinus rhythm for 12 hours after cardiac surgery. BUN = blood urea nitrogen; beta = beta-blocker use; Ca = calcium level; CRP = C-reactive protein; Digoxin = digoxin use; EF = ejection fraction; Glu = glucose level; HR = heart rate; IVS = interventricular septum thickness; K = potassium level; LAAD = left atrial anteroposterior diameter; LVED = left ventricular end-diastolic dimension; Na = sodium level; NT = N-terminal pro-B-type natriuretic peptide; PAP = pulmonary artery pressure; RAM = right atrial maximum diameter; RVAD = right ventricular anteroposterior diameter; Cr = creatinine level.

## 4. Discussion

AF is the most common sustained arrhythmia in patients undergoing cardiac surgery, and preoperative persistent AF significantly increases the risk of hemodynamic instability, myocardial dysfunction, and adverse postoperative outcomes^12–14^. This retrospective cohort study evaluates the efficacy of intraoperative intravenous amiodarone administration during rewarming in CPB for patients with preoperative persistent AF, and develops a predictive model for AF recurrence within 12 hours after cardioversion. Our research findings indicate that administering 150 mg of amiodarone via an oxygenator during the initial phase of CPB rewarming significantly increases the success rate of maintaining sinus rhythm within 12 hours postoperatively (52.5% vs. 7.8%). Additionally, we identified that amiodarone use is associated with improved rhythm control, and we identify six independent predictors of sustained sinus rhythm maintenance: age, LAAD, LVED, RVAD, serum Ca, and comorbidity burden (Grade 3 vs. Grade 1). The predictive model shows strong discriminative performance, with an area under the receiver operating characteristic curve (AUC) of 0.866 in the training set and 0.795 in the validation set, supporting its potential clinical utility.

This finding has important clinical implications. Patients with preoperative persistent AF undergoing cardiac surgery often experience a 20–30% reduction in cardiac output due to loss of atrial mechanical function, irregular ventricular rate, and atrioventricular dyssynchrony, significantly increasing the risk of perioperative hemodynamic instability^15^. Although CPB temporarily supports circulation, the early reperfusion phase following aortic unclamping represents one of the most electrophysiologically vulnerable periods, during which malignant arrhythmias or persistent/recurrent AF are common. Administering a 150 mg intravenous bolus of amiodarone via the CPB oxygenator at the initiation of systemic rewarming aligns precisely with this critical "therapeutic window." Amiodarone, a multi-channel blocking agent, is classified as a class III antiarrhythmic drug but also exhibits additional effects including sodium ^16^, and calcium channel blockade ^17, 18^, as well as non-competitive α- and β-adrenergic receptor inhibition ^19^. It stabilizes the myocardial electrophysiological environment by prolonging the action potential duration and effective refractory period, thereby suppressing re-entrant circuits and triggered activity, promoting sinus rhythm restoration and reducing early recurrence ^20^.Studies have shown that intravenous amiodarone during the perioperative period achieves higher success rates compared to other antiarrhythmic agents, particularly when administered prior to electrical cardioversion (ECV), which significantly reduces the immediate recurrence of AF ^21^. Our study extends these findings by precisely timing the intervention at the onset of rewarming and defining success as sustained sinus rhythm within 12 hours post-cardioversion—a clinically relevant window for hemodynamic stability and end-organ perfusion. Our results demonstrate that this strategy enables over half of patients to maintain stable sinus rhythm (52.5% vs. 7.8%), significantly outperforming the non-intervention group, suggesting that the rewarming phase represents an optimal opportunity for pharmacological rhythm control.

Notably, although the A group achieved superior rhythm control, they exhibited significantly higher NT-proBNP levels on postoperative days 2 and 3, suggesting potentially increased ventricular wall stress or neurohormonal activation. This may be attributed to post-cardioversion atrial stunning, autonomic imbalance, or amiodarone-related negative inotropic effects. In contrast, the NI group, despite minimal sinus conversion, showed markedly elevated myocardial injury markers (cTnI), stronger early inflammatory responses (NEUT), higher BUN levels, greater reliance on MIL, and increased need for HD, ultimately resulting in prolonged LOHS. These findings suggest that maintaining AF without active rhythm control may impose a greater systemic organ stress burden, whereas amiodarone-mediated rhythm stabilization, despite potential compensatory demands (e.g., increased norepinephrine use), may overall reduce the risk of multi-organ dysfunction.

In the multivariable logistic regression model, age emerged as a significant negative predictor of successful cardioversion (OR = 0.478, 95% CI: 0.288–0.792). This is consistent with age-related atrial fibrosis, ion channel remodeling, and autonomic dysfunction, all of which contribute to AF persistence^22–24^. Each additional year of age was associated with a 52% reduction in the odds of maintaining sinus rhythm, highlighting the challenge of rhythm control in older patients.

Structural remodeling of the atria and ventricles also played a pivotal role^25, 26^. LAAD was inversely associated with success (OR = 0.520, 95% CI: 0.327–0.827), indicating that each millimeter increase in left atrial size reduced the likelihood of rhythm stabilization by approximately 48%. This reinforces the concept that left atrial enlargement reflects advanced atrial myopathy and serves as a substrate for re-entry. Similarly, LVED (OR = 0.572, 95% CI: 0.385–0.851) and RVAD (OR = 0.667, 95% CI: 0.488–0.911) were independent negative predictors, suggesting that biventricular dilation contributes to global electrical instability, possibly through interatrial conduction delay or mechanical stretch-induced ectopy^27^.

Serum calcium concentration was another significant predictor (OR = 0.724, 95% CI: 0.548–0.955), with higher levels linked to lower success rates. While hypocalcemia is traditionally associated with arrhythmogenesis, our finding suggests that hypercalcemia—or a relative excess in the perioperative setting—may promote calcium overload during reperfusion, triggering early afterdepolarizations and increasing AF vulnerability. This paradoxical association warrants further investigation in larger cohorts.

The strongest predictor was comorbidity burden, with patients classified as Grade 3 having an 88% lower odds of maintaining sinus rhythm compared to Grade 1 (OR = 0.119, 95% CI: 0.048–0.294). This variable likely reflects the cumulative impact of systemic inflammation^28^, endothelial dysfunction^29^, and neurohormonal activation^30^ on atrial electrophysiology. Conditions such as chronic kidney disease^31^, diabetes^32^, and obstructive sleep apnea^33^ may synergistically exacerbate atrial remodeling and reduce responsiveness to antiarrhythmic drugs.

Notably, several conventional markers—including LVEF, RAM, PAP, and post-CPB potassium (K)—did not retain statistical significance in the final model. While PAP showed a trend toward association (OR = 0.680, 95% CI: 0.457–1.012), it did not reach significance, possibly due to collinearity with RVAD or effective intraoperative management. The lack of association with LVEF suggests that global systolic function may be less critical than structural dimensions in determining arrhythmia susceptibility in this population. Similarly, although hypokalemia is a known trigger for AF, routine correction in the ICU may have minimized its independent effect.

This study has several strengths. First, we applied IPTW with weight truncation to balance baseline characteristics between the amiodarone and non-intervention groups, reducing selection bias and enhancing comparability in this observational design. Second, the model was developed in a training cohort and validated in an independent subset, demonstrating good discriminative ability in both sets. The resulting nomogram provides a user-friendly tool for clinicians to estimate individual recurrence risk and guide prophylactic or therapeutic decisions.

Nonetheless, limitations should be acknowledged. As a single-center retrospective study, unmeasured confounders—such as genetic predisposition, autonomic tone, or subtle differences in surgical technique—may influence outcomes. The absence of long-term follow-up limits assessment of rhythm durability and hard endpoints such as stroke or mortality. Additionally, postoperative antiarrhythmic management was not standardized, which may introduce variability in care. Importantly, the amiodarone dosing regimen in this study was fixed at 150 mg intravenously during CPB rewarming, without adjustment for key patient-specific factors such as body weight, age-related changes in hepatic metabolism, or underlying cardiac function. This lack of individualization may have led to subtherapeutic concentrations in some patients—potentially underestimating the drug’s true efficacy—while resulting in supratherapeutic levels in others, increasing the risk of adverse effects such as hypotension and bradycardia. These pharmacokinetic variations could have confounded both the safety profile and the apparent treatment effect, highlighting the need for weight- or clearance-adjusted dosing strategies in future investigations. Prospective, randomized trials are therefore needed to confirm the benefit of this amiodarone protocol and to evaluate whether risk-stratified, personalized dosing—guided by clinical predictors such as those in our model—can optimize both rhythm control and clinical safety.

## 5. Conclusion

In patients with preoperative AF undergoing cardiac surgery, amiodarone administration during rewarming significantly improves early rhythm stabilization within 12 hours. Although the A group had transiently elevated NT-proBNP, the NI group exhibited greater myocardial injury (↑cTnI), stronger inflammation (↑NEUT), higher BUN, increased MIL and HD use, and longer LOHS, indicating greater systemic organ stress. Our predictive model—incorporating age, LAAD, LVED, RVAD, serum calcium, and comorbidity grade—showed good discrimination for early AF recurrence in both training and validation cohorts. The corresponding nomogram may aid clinical risk stratification. These results support proactive amiodarone use during rewarming as an effective strategy to improve short-term rhythm outcomes. Overall, proactive amiodarone administration during rewarming is an effective strategy for improving short-term rhythm outcomes. These results support its role in perioperative AF management and lay the groundwork for future risk-guided or individualized antiarrhythmic approaches in this high-risk population.

## CRediT authorship contribution statement

All authors contributed significantly to this work and supported the publication of the manuscript. **Liangcheng Zhang**: conceptualization. **Linlin Chen** and **Qi Gao**: writing - original draft preparation. **Liangcheng Zhang**: editing, and revising. **Liangcheng Zhang**: supervision.

## Data Availability

The datasets generated during and/or analyzed during the current study are available from the corresponding author on reasonable request.

